# Fructosamine and Triglycerides - Developing an Index for Assessing Insulin Resistance

**DOI:** 10.1101/2024.05.19.24307591

**Authors:** Luís Jesuíno de Oliveira Andrade, Gabriela Correia Matos de Oliveira, Luísa Correia Matos de Oliveira, Luiz Felipe Moreno de Brito, Luís Matos de Oliveira

## Abstract

**Introduction:** Insulin resistance (IR) is a metabolic condition where cells become less responsive to insulin, affecting glucose absorption and leading to diseases like type 2 diabetes. Factors like obesity and high triglycerides worsen IR. Fructosamine is a key marker for short-term glycemic control and can indicate IR. Various methods, like the euglycemic clamp and HOMA-IR index, assess IR, with the TyG index emerging as a simple and reliable tool due to its strong correlation with IR.

**Objective:** To develop and evaluate a novel IR index that incorporates both fructosamine and triglyceride levels, aiming to improve the accuracy of IR assessment compared to existing methods.

**Methods:** This study investigated IR using established methods and proposed a novel TrigFruc index incorporating fructosamine. Data from 200 individuals was analyzed to assess the TrigFruc index’s performance compared to existing methods (HOMA-IR and TyG index). Descriptive statistics summarized participant demographics, and Pearson’s correlation evaluated the relationship between TrigFruc and HOMA-IR and between TrigFruc and TyG index. A receiver operating characteristic (ROC) analysis identified the optimal TrigFruc cut-off point for IR detection, comparing its sensitivity and specificity to HOMA-IR and TyG index.

**Results:** Study with 200 participants (63% female, avg. age 46.6 years), the established HOMA-IR index identified IR in 32% (64 individuals) while the TyG index found it in 66% (132). The new TrigFruc index showed a weaker correlation with HOMA-IR (r=0.28) compared to the TyG index (r=0.44). For detecting IR based on HOMA-IR, the optimal TrigFruc cut-off (Ln 4.57) had just 50% sensitivity (correctly identifying half of true IR cases) but low specificity (23%, high misclassification of non-IR cases). When compared to the TyG index, the optimal TrigFruc cut-off (Ln 4.74) showed excellent performance with high sensitivity (85%, strong ability to identify true IR) and specificity (95%, low misclassification of non-IR).

**Conclusion:** The TrigFruc index, incorporating fructosamine, shows promise for IR assessment. Compared to HOMA-IR, it exhibits better correlation with the TyG index and superior sensitivity and specificity for IR detection when using the TyG index as a reference.

## INTRODUCTION

Insulin resistance (IR) is a metabolic condition where cells become less responsive to the effects of insulin. This leads to a reduced ability to absorb glucose from the bloodstream.^1^ IR is a major contributor to various diseases, including metabolic syndrome, type 2 diabetes (T2DM), and heart problems.^2^ Accurately assessing IR is critical for early detection and effective treatment of these conditions.

Several factors, both genetic and environmental, contribute to IR. It’s often linked to obesity, physical inactivity, and unhealthy eating habits. The core issue behind IR is a malfunctioning insulin receptor signaling pathway, leading to decreased glucose uptake and utilization by muscles and fat tissues.^3^

Studies have shown that triglycerides significantly impact IR.^4^ High triglyceride levels contribute to a buildup of fatty acids in non-fatty tissues, especially muscle and liver. This abnormal fat storage disrupts insulin signaling and worsens IR.^5^

Fructosamine, a marker of average blood sugar levels over the past 2-3 weeks, offers valuable insight into short-term glycemic control.^6^ It reflects the effectiveness of diabetes management.^7^ Research suggests a strong link between elevated fructosamine and IR, indicating its potential as a reliable indicator of IR.^8^

The gold standard for directly measuring IR is the euglycemic clamp technique.^9^ Another widely used tool for assessing IR in individuals is the homeostasis model assessment for IR (HOMA-IR) index.^10^

In recent years, a new index called the Triglyceride-Glucose (TyG) index has emerged as a promising tool for assessing insulin resistance (IR) due to its simplicity and reliability.^11^ Multiple studies have confirmed the strong correlation between the TyG index and IR, highlighting its potential to identify individuals at risk for developing diabetes.^12^

The TyG index leverages the combined power of triglyceride and glucose levels, both of which are critical factors in IR. Elevated triglycerides indicate abnormal lipid metabolism, a common characteristic in individuals with IR. Similarly, high blood sugar levels reflect impaired glucose metabolism, another hallmark of IR. By incorporating both these parameters, the TyG index provides a comprehensive picture of IR, making it a valuable asset for both clinical practice and research.

This study proposes the development of a IR index, matching triglyceride measurements with an estimate of blood sugar levels using fructosamine, creating a more comprehensive and accurate measure of IR through this combined approach. This new index would have the potential to significantly improve the diagnosis and management of various conditions associated with IR.

## METHOD

This investigation employed the HOMA-IR index to assess IR. The formula for HOMA-IR involved multiplying fasting insulin concentration (in μU/mL) by fasting blood glucose level (in mg/dL) and then dividing the product by a conversion factor (415): Fasting insulin in betaU/mL × fasting glucose in mg/dL) / 415.^13^ Consistent with prior studies, a HOMA-IR value greater than or equal to 3.4 was considered indicative of IR. This threshold aligns with the gold standard hyperglycemic-hyperinsulinemic clamp method and offers optimal prediction for the development of diabetes.

Furthermore, the TyG index was utilized calculated as the natural logarithm of the product obtained by multiplying fasting triglycerides (mg/dL) by fasting glucose (mg/dL) and then dividing by 2: Ln[fasting triglycerides (mg/dL) fasting glucose (mg/dL)/2]. The TyG index is expressed on a logarithmic scale, and a value of 4.49 was established as the benchmark for identifying IR.^14^

To leverage fructosamine as a marker of glycemic control, a novel triglyceride-fructosamine (TrigFruc) index was developed. This index adopted the same formula as the TyG index, but instead of using fasting glucose, it incorporated an estimated average glucose value derived from fructosamine concentration: Ln[fasting triglycerides (mg/dL) ((0.5157 x Fructosamine) – 20)/2].^15^

### Data Acquisition

This study involved the analysis of data from 200 individuals. The data was obtained by retrospectively examining existing laboratory records. Demographic information, along with biochemical measurements, was included when complete data sets were available. These measurements encompassed sex, age, fasting glucose levels, triglyceride concentrations, insulin levels, HOMA index values, TyG index values, fructosamine levels, and estimated average glucose derived from fructosamine measurements.

### Data Analysis

An overview of the participants’ demographic characteristics was generated using descriptive statistics. For continuous variables, such as fasting glucose or age, the results are presented as the average value along with the standard deviation to depict the range of data. Categorical variables, like gender, are expressed as the number of individuals within each category and the corresponding percentage.

The strength and direction of the relationship between the triglycerides-fructosamine index and the HOMA-IR index were evaluated using Pearson’s correlation coefficient.

To identify the optimal cut-off point for the triglycerides-fructosamine index in identifying IR, a receiver operating characteristic (ROC) analysis was performed. This analysis compared the triglycerides-fructosamine index to the established HOMA-IR index and the TyG index. The ROC analysis yielded information on the sensitivity, specificity, positive and negative predictive values, as well as positive and negative likelihood ratios of the triglycerides-fructosamine index.

### Research Ethics

This research adhered to the ethical principles outlined in the Declaration of Helsinki. The project received approval from the National Commission for Research Ethics in Brazil (CONEP), with a registry number of 2,464,513. To safeguard patient privacy, we implemented rigorous protocols for data confidentiality and protection.

## RESULTS

The analysis included data from 200 subjects. Among these participants, 126 (63.0%) were female and 74 (37.0%) were male. The average age was 46.56 years, with a standard deviation of 18.98 years. Insulin resistance (IR) was detected in 64 (32.0%) individuals using the HOMA-IR index, while the TyG index identified IR in 132 (66.0%) participants. Notably, the HOMA-IR and TyG indices did not yield consistent results for the same individuals. A more detailed breakdown of the demographic characteristics is provided in Table 1.

**Table 1.**
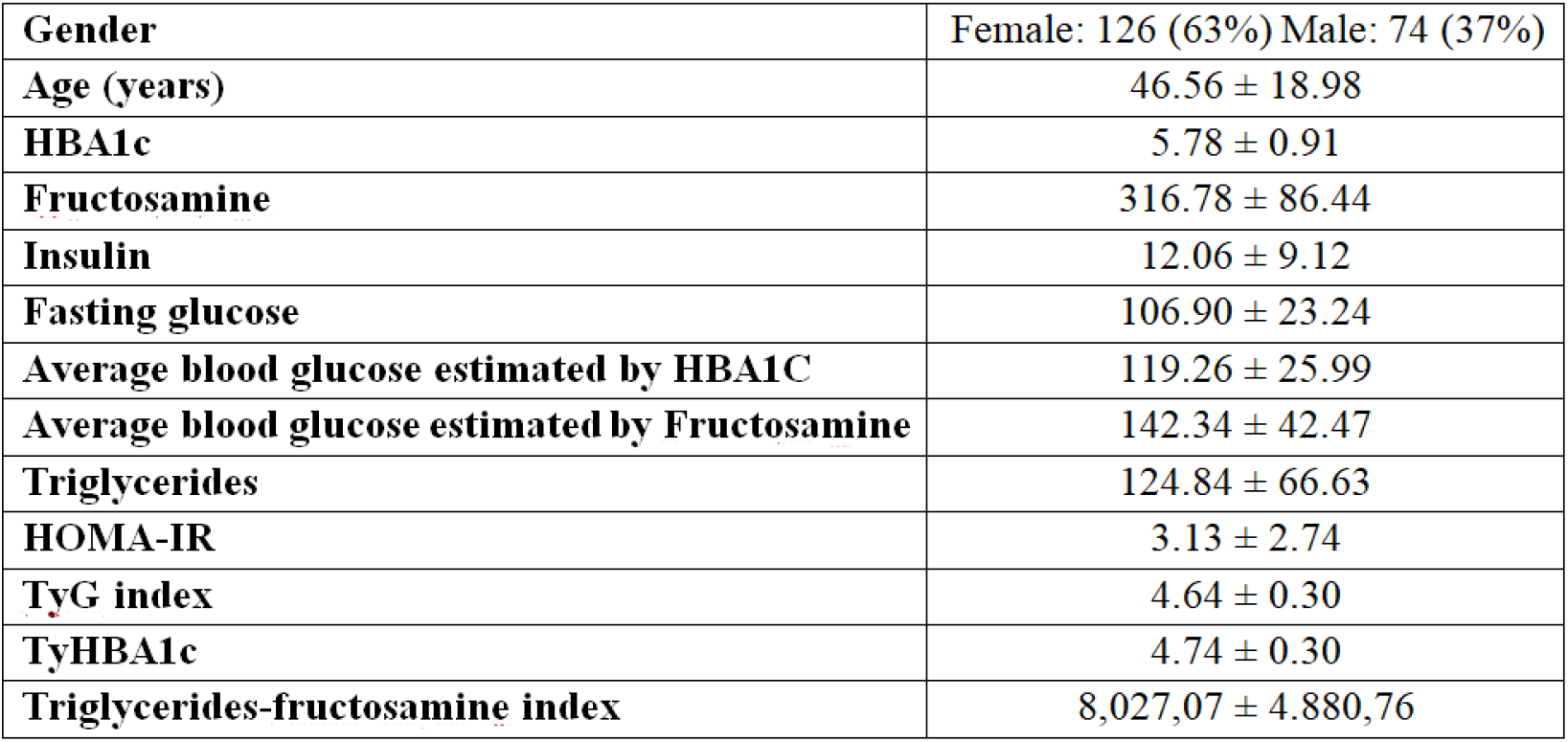
Demographis characteristics.

|  |  |
| --- | --- |
| **Gender** | Female: 126 (63%) Male: 74 (37%) |
| **Age (years)** | 46.56 ± 18.98 |
| **HBA1c** | 5.78 ± 0.91 |
| **Fructosamine** | 316.78 ± 86.44 |
| **Insulin** | 12.06 ± 9.12 |
| **Fasting glucose** | 106.90 ± 23.24 |
| **Average blood glucose estimated by HBA1C** | 119.26 ± 25.99 |
| **Average blood glucose estimated by Fructosamine** | 142.34 ± 42.47 |
| **Triglycerides** | 124.84 ± 66.63 |
| **HOMA-IR** | 3.13 ± 2.74 |
| **TyG index** | 4.64 ± 0.30 |
| **TyHBA1c** | 4.74 ± 0.30 |
| **Triglycerides-fructosamine index** | 8,027,07 ± 4.880,76 |

**Table 2.** Correlation between the TrigFruc index and HOMA-IR.

| Correlations |  |  |  |
| --- | --- | --- | --- |
|  |  | TYGFRUT | HOMA-IR |
| TYGFRUT | Pearson Correlation | 1,000 | ,280 |
|  | Sig. (2-tailed) |  | ,000 |
|  | N | 200 | 200 |
| HOMA-IR | Pearson Correlation | ,280 | 1,000 |
|  | Sig. (2-tailed) | ,000 |  |
|  | N | 200 | 200 |

Our findings indicate a weaker correlation between the TrigFruc index and the HOMA-IR index compared to the TrigFruc index and the TyG index. The Pearson correlation coefficient between the TrigFruc index and the HOMA-IR index was *r* = 0.280 (Table 1), whereas the Pearson correlation coefficient between the TrigFruc index and the TyG index was *r* = 0.441 (Table 3).

**Table 3.** Correlation between the TrigFruc index and TyG index.

| Correlations |  |  |  |
| --- | --- | --- | --- |
|  |  | TYG INDICE | TYGFRUT |
| TYG INDICE | Pearson Correlation | 1,000 | ,441<sub>a</sub> |
|  | Sig. (2-tailed) |  | ,000 |
|  | N | 200 | 200 |
| TYGFRUT | Pearson Correlation | ,441<sub>a</sub> | 1,000 |
|  | Sig. (2-tailed) | ,000 |  |
|  | N | 200 | 200 |

Analysis of the ROC curve revealed that a logarithmic transformed TrigFruc index value of 4.57 yielded the optimal cut-off point for distinguishing between IR cases based on the HOMA-IR index. This cut-off value achieved a sensitivity of 50.0%, indicating it correctly identified half of the true IR cases, but a lower specificity of 23.0%, meaning a significant portion of non-IR cases were misclassified (Figure 1).

**Figure 1.**
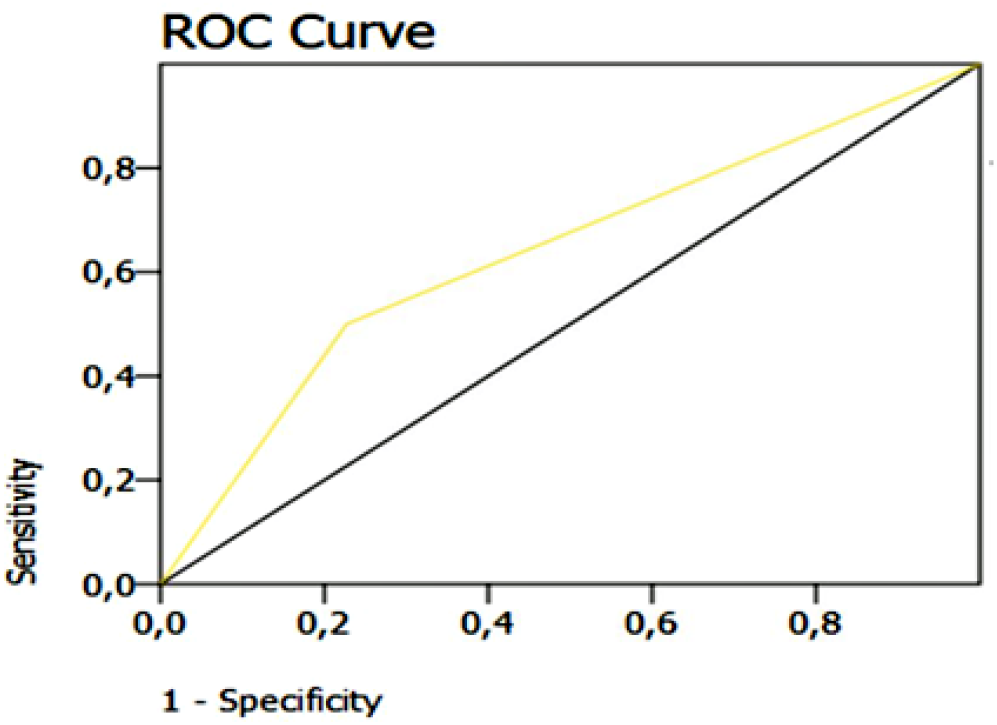
ROC analysis: TrigFruc index compared to the HOMA-IR index

As evident from the ROC curve analysis (Figure 2), the optimal cut-off point for the TrigFruc index in identifying IR relative to the TyG index was Ln 4.74. This cut-off exhibited high sensitivity (85.0%) signifying a strong ability to correctly classify true IR cases. Additionally, it demonstrated specificity of 95.0%, indicating a low rate of misclassifying non-IR cases.

**Figure 2.**
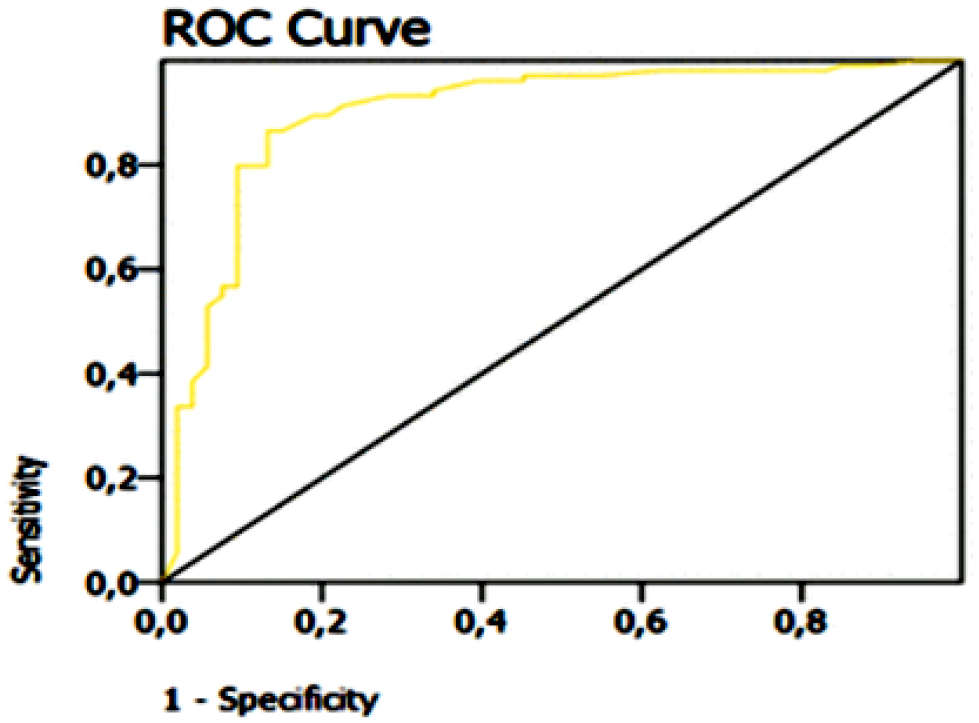
ROC analysis: TrigFruc index compared to the TyG index

## DISCUSSION

This research investigated whether a new indicator, calculated by multiplying triglyceride levels by the estimated average glucose derived from fructosamine, could serve as an alternative to the TyG index for assessing insulin resistance. The study compared the effectiveness of this new index, called the TrigFruc index, to the TyG index. In medical research, sensitivity refers to a test’s accuracy in identifying people with a specific condition. Specificity, on the other hand, indicates how well a test can rule out the condition in healthy individuals. Sensitivity reflects the proportion of true positives, while specificity represents the proportion of true negatives. These measures are essential for determining the trustworthiness and validity of diagnostic tests used in everyday clinical practice.^16,17^

In our investigation, the TrigFruc index, when compared to the HOMA-IR index for insulin resistance assessment, demonstrated moderate sensitivity and low specificity. However, the extensively utilized HOMA-IR index exhibits remarkable sensitivity and specificity in gauging insulin resistance risk, even when compared to more intricate methodologies.^18^

Current clinical practice relies on indices like HOMA-IR and TyG to evaluate IR. While HOMA-IR focuses on the interplay between fasting glucose and insulin, the TyG index incorporates triglycerides but omits insulin.^13,14^ Our research investigated whether substituting fructosamine-derived estimated average glucose for fasting glucose in the TyG index formula could lead to more precise IR assessments. This approach hinges on the superior stability of fructosamine, reflecting average glycemic levels over a 2-3 week window compared to a single fasting glucose measurement. Thus, our study compared the performance of the newly proposed TrigFruc index to the established HOMA-IR and TyG indices in a cohort with varying degrees of IR.

A growing body of evidence underscores the positive correlation between the TyG index and both the HOMA-IR index and the hyperinsulinemic-euglycemic clamp test. This has propelled the TyG index to prominence and readily calculable surrogate marker for IR.^19^ Our present study aimed to investigate the association between the novel TrigFruc index, proposed by the authors, and the well-established HOMA-IR index. While Pearson’s correlation analysis revealed a statistically significant positive association between the two indices, the observed strength of this relationship was demonstrably lower compared to the correlation between the TrigFruc index and the TyG index. This finding suggests that the TrigFruc index may offer a more robust and reliable measure of IR compared to the HOMA-IR index. The potential advantage of the TrigFruc index likely stems from its inclusion of fructosamine. Unlike the HOMA-IR index, which relies solely on potentially variable fasting glucose and insulin levels, the TrigFruc index incorporates fructosamine. This glycated protein offers a stable marker of average glycemia over a 2-3 weeks period, mitigating the influence of transient fluctuations.^20^

Our investigation into the novel TrigFruc index, a marker for insulin resistance, revealed a robust correlation with established indices like the TyG and HOMA-IR. This innovative approach leverages fructosamine to estimate average blood glucose levels over a 2-3 week window, enhancing the precision of IR assessment compared to methods reliant on single fasting glucose measurements. These findings suggest the potential utility of the TrigFruc index in clinical practice. Its ability to capture a more comprehensive picture of glycemic control could contribute to earlier diagnosis, improved monitoring, and potentially more effective management of IR-related conditions.

## CONCLUSION

This study explored the TrigFruc index, as a novel marker for IR incorporating fructosamine. While the TrigFruc index exhibited a weaker correlation with the established HOMA-IR index, it demonstrated a stronger association with the TyG index. Notably, the TrigFruc index achieved high sensitivity and specificity for IR detection when compared to the TyG index. These findings suggest the TrigFruc index holds promise as a accurate and reliable tool for IR assessment, particularly when used in conjunction with the TyG index.

## Data Availability

All data produced in the present work are contained in the manuscript

